# Risk factors for West Nile neuroinvasive disease and mortality in the United States, 2013-2024

**DOI:** 10.1101/2025.07.21.25331933

**Authors:** Seth D. Judson, David Dowdy

## Abstract

**Importance:** West Nile virus (WNV) is the leading mosquito-borne infection in the United States and can cause West Nile fever (WNF) or West Nile neuroinvasive disease (WNND), resulting in substantial morbidity and mortality. Contemporary risk factor analyses are needed to identify populations at increased risk for severe outcomes and target interventions accordingly.

**Objective:** To assess risk factors for WNND and mortality among adults with WNV infection using a large national cohort from federated real-world data.

**Design:** A retrospective cohort analysis from 2013-2024, consisting of de-identified electronic health record data from the TriNetX Research Network, including adult patients with an ICD-10 diagnosis consistent with WNV infection. Data were accessed from March 28^th^ to April 30^th^, 2025.

**Setting:** The retrospective cohort included patients from 65 healthcare organizations from across the United States, using the TriNetX Research Network.

**Participants:** The overall initial cohort included 3,064 adult patients with an ICD-10 diagnosis consistent with WNV infection, including 1,328 with WNF and 1,206 with WNND.

**Exposures:** Demographic characteristics (age, sex, race, ethnicity), comorbidities (hematologic malignancy, ischemic heart disease, other forms of heart disease, diabetes, HIV, chronic kidney disease (CKD), liver disease, hypertension, alcohol related disorders, cerebrovascular disease (CEVD), chronic obstructive pulmonary disease, asthma, multiple sclerosis, dementia, rheumatoid arthritis, organ transplant), and medications (immune suppressants, antineoplastics).

**Main Outcomes and Measures:** The primary outcomes were the development of WNND and all-cause mortality (30-day, 90-day, and overall mortality).

**Results:** Among all patients diagnosed with WNV infection, those with WNND were older (mean age 59 vs 55, p-value <0.0001) and more often male (61% vs 48%, p-value <0.0001). Significant risk factors for WNND included age (adjusted hazard ratio [aHR] 1.11 per decade, 95% confidence interval [CI] 1.06-1.49), male sex (aHR 1.29, 1.15-1.45), CKD (aHR 1.21, 1.002-1.45), CEVD (aHR 1.22, 1.03-1.45), hematologic malignancy (aHR 1.38, 1.09-1.76), immune suppressant use (aHR 1.43, 1.11-1.83), hypertension (aHR 1.18, 1.04-1.34), alcohol related disorders (aHR 1.54, 1.20-1.97), and multiple sclerosis (aHR 2.3, 1.62-3.37). Significant risk factors for mortality were WNND (aHR 2.49 for 30-day mortality, 95% CI 1.37-4.52), age (aHR 1.32 per decade, 95% CI 1.07-1.60), CKD (aHR 2.08, 95% CI 1.01-3.93), and CEVD (aHR 2.00, 95% CI 1.14-3.50).

**Conclusions and Relevance:** Risk factors for WNND broadly reflect impaired immune response and/or central nervous system vulnerability. Patients who developed WNND were at substantially increased risk of death. Targeted prevention strategies and countermeasures for those at greatest risk for WNND could substantially reduce morbidity and mortality.

**Key Points:** *Question:* What are risk factors for West Nile virus neuroinvasive disease and mortality in the United States between 2013 and 2024?

*Findings:* In this retrospective cohort analysis, we found that increased age, chronic kidney disease, and cerebrovascular disease are significantly associated with both West Nile neuroinvasive disease and mortality. Additionally, male sex, hematologic malignancy, immune suppressants, hypertension, alcohol related disorders, and multiple sclerosis were significant risk factors for West Nile neuroinvasive disease.

*Meaning:* An aging population in the United States with increasing rates of comorbidities and immunosuppression faces an increased risk for severe disease from West Nile virus infection.

## Introduction

West Nile virus (WNV) first emerged in the United States (U.S) in 1999 and is an ongoing threat, causing more than 80% of all cases of mosquito-borne infections in the U.S annually.^1^ Now endemic throughout most of the continental U.S, WNV is transmitted from bird reservoir hosts to humans primarily via *Culex* mosquitoes.^2^ It is estimated that approximately 75% of people infected with WNV are asymptomatic, while 25% develop a non-specific febrile syndrome called West Nile fever (WNF).^3–5^ A subset of WNV infections (<1%) can progress to West Nile neuroinvasive disease (WNND), which has a case fatality of ∼10% and can cause lifelong neurological complications.^6,7^

One of the priorities for the 2024 U.S Centers for Disease Control and Prevention (CDC) National Strategy for Vector-borne Diseases is to reduce the annual number of WNND cases to below 500 by 2035.^1^ To achieve this goal, a better understanding is needed of the risk factors associated with severe WNV infection, including WNND and mortality. Unfortunately, the majority of risk factor analyses for severe WNV infection in the U.S were performed over a decade ago and are limited in their geographic and/or population scope.^8–10^ One of the largest risk factor analyses covering multiple regions in the U.S was a retrospective cohort from 2008 to 2010 using CDC ArboNet data from 19 jurisdictions from primarily western and midwestern states and included 1090 adult WNV cases (641 with WNND).^9^ Additionally, comparisons of risk factors have been limited by study heterogeneity and small sample sizes, as noted by both a recent scoping review and a systematic review/meta-analysis.^8,10^ Given changing demographics and increasing prevalence of comorbidities, such as immunosuppression, in the U.S., there is a need to reassess risk factors for WNND and disease severity.^11^

Recent advancements in research networks and the electronic medical record (EMR) over the past decade provide new opportunities to analyze additional data in the context of WNV infection. In particular, federated learning provides the opportunity to analyze real-world infectious disease data from multiple clinical sources^12^, significantly increasing sample sizes and geographic representation. To address gaps in our existing knowledge regarding risk factors for WNV, we used the TriNetX Research Network,^13^ a federated real-word data and analytics platform from a large research network of healthcare organizations (HCOs), to identify factors associated with WNND and mortality across the U.S. from 2013 to 2024.

## Methods

### Ethical Considerations

The retrospective cohort study from TriNetX Research Network included only de-identified aggregate data and does not require IRB approval. We used the Strengthening the Reporting of Observational Studies in Epidemiology (STROBE) cohort checklist for reporting.

### Cohort identification

We identified retrospective cohorts from querying TriNetX Research Network, a federated real-world data and analytics platform containing data from over 100 HCOs across the U.S. Using *International Classification of Diseases and Related Health Problems, Tenth Revision (ICD-10)* codes, we formed multiple cohorts of de-identified and aggregated data from adult patients (≥18 years of age) diagnosed with WNV infection from 2013-2024 (S Table 1). We used standardized ICD-10 codes for WNF (A92.30, A92.39) and WNND (A92.31, A92.32).

Data from the TriNetX Research Network were accessed from March 28^th^ to April 30^th^. We also compared the patient counts to national CDC ArboNet data from 2013-2024.^14^

### Risk factor identification

To identify potential risk factors for WNND and mortality, we conducted a literature review in PubMed using the search terms “West Nile” and “risk”. We also referenced a scoping review from 2017 and systematic review from 2025 to identify demographic and clinical risk factors that have been studied in association with WNV outcomes.^8,10^ We selected risk factors that previously have been assessed or associated with WNND, severity, and/or mortality. Our final list of potential risk factors included age, sex, ethnicity, race, 16 co-morbidities, and five classes of medications (S Table 2). For our analysis, we included risk factors that were present within a 30 day-period before or on the same day as WNV diagnosis (details in Supplementary Material).

### Statistical Analysis

Descriptive statistics of the TriNetX cohorts were calculated in TriNetX, which compares continuous data using independent t-tests and categorical data using ^2^ or Fisher’s exact tests. We selected potential risk factors that were significant in bivariate analyses (two-sided p-value <0.05) to use in our multivariable analyses, with the exceptions of subgroups of immune suppressants (tacrolimus and mycophenolate mofetil [MMF]) and solid organ transplant, which both had a high level of overlap with the broader category of immune suppressant use.

The primary outcomes for our analysis were WNND and all-cause mortality. Given that the analytical functionality of TriNetX is geared towards time-to-event analysis, we used multivariable Cox proportional hazard models for our primary analyses. Since mortality associated with WNV may vary by time period, we compared overall mortality as well as acute mortality (within 30 days and within 90 days). TriNetX lacks functionality to test the proportionality assumption in multivariable models, but can test this assumption in univariable analysis with categorial covariates. Therefore, we checked the proportionality assumption through two mechanisms. First, we conducted univariable analysis with each categorial covariate that was significantly associated with WNND and used Schoenfield residuals to evaluate proportionality (performed in TriNetX using a chi-squared test; a p-value >0.05 suggests the proportionality assumption likely holds for that covariate). For mortality, we also compared HRs across different time segments (overall, 30 days, and 90 days).

## Results

### West Nile Patient Characteristics 2013-2024

From 2013 through 2024, 3,064 adults were diagnosed with WNV infection (including WNND) from 65 HCOs in the TriNetX Research Network. Of these patients, 1,328 (43%) had ICD-10 codes consistent with WNF and 1,206 (39%) had codes consistent with WNND. The distribution of these patients by geographic region included 1053 (35%) in the West, 1052 (35%) in the South, 459 (15%) in the Midwest, and 379 (13%) in the Northeast. Baseline characteristics of the cohorts stratified by WNND are shown in Table 1 and by mortality in S Table 3.

**Table 1.**
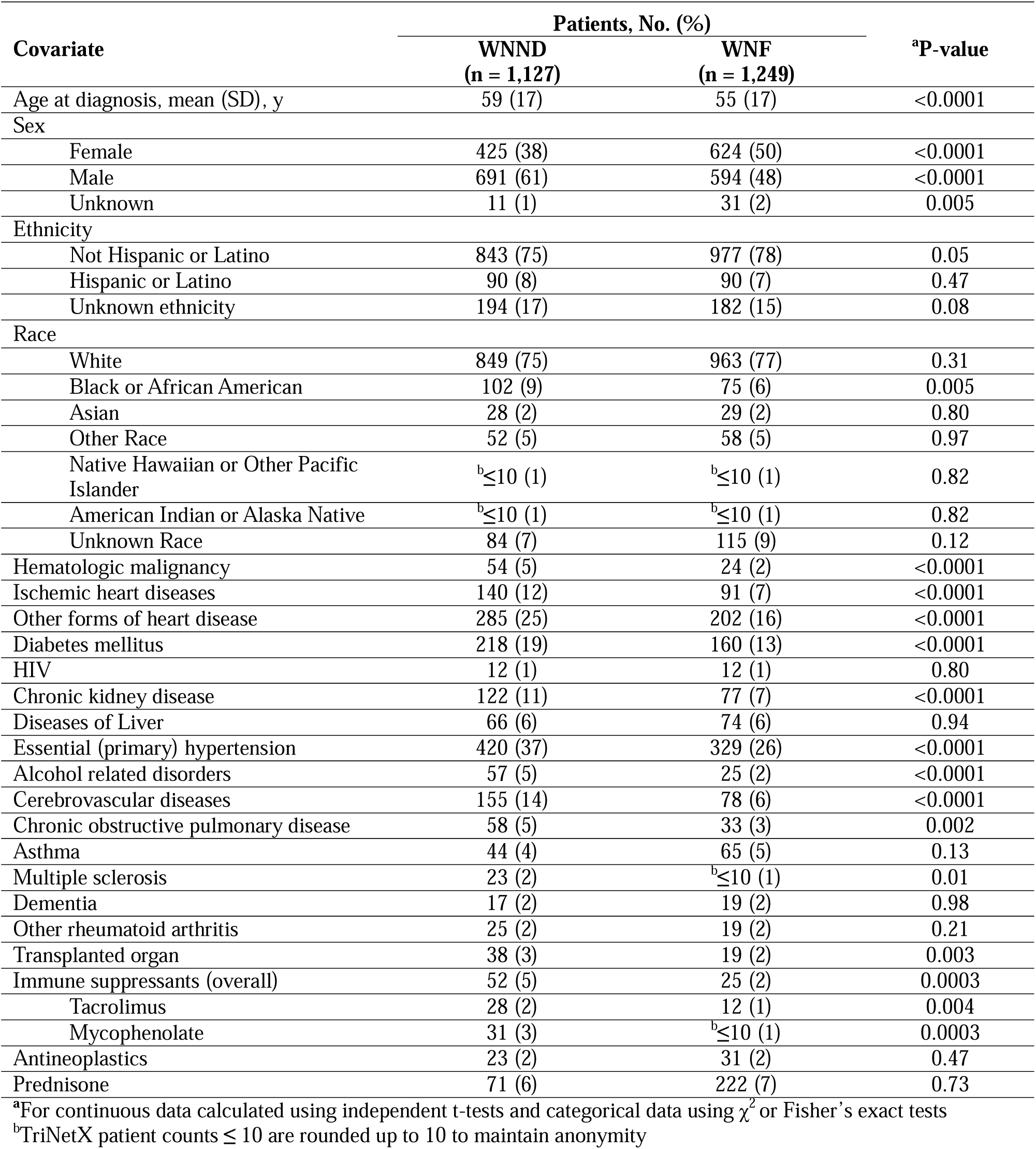
Baseline Characteristics of Patients with West Nile Neuroinvasive Disease (WNND) and West Nile Fever (WNF), United States, 2013-2024.

Compared to CDC ArboNet data, the TriNetX cohort represented 19% (3,064/16,413) of nationally reported WNV cases and 37% (1,206/3,258) of nationally reported WNND cases from 2013 through 2024.

### Risk factors for WNND

Patient-level characteristics that were significantly associated with WNND in bivariate analysis included age; male sex; Black or African American race; ischemic heart disease; other heart disease; malignant neoplasms of lymphoid, hematopoietic and related tissue (hematologic malignancy); diabetes; CKD; hypertension; alcohol related disorders; chronic obstructive pulmonary disease (COPD); cerebrovascular disease (CEVD); multiple sclerosis (MS); solid organ transplant; and immune suppressant use (including subgroups of tacrolimus and MMF).

In multivariable analysis, risk factors significantly associated with WNND included age (per decade) (aHR 1.11, 95% CI 1.06-1.49), male sex (aHR 1.29, 1.15-1.45), CKD (aHR 1.21, 1.002-1.45), CEVD (aHR 1.22, 1.03-1.45), hematologic malignancy (aHR 1.38, 1.09-1.76), immune suppressant use (aHR 1.43, 1.11-1.83), hypertension (aHR 1.18, 1.04-1.34), alcohol related disorders (aHR 1.54, 1.20-1.97), and MS (aHR 2.3, 1.62-3.37) (Table 2). The proportionality assumption was met for sex (*χ*^2^=0.41, p = 0.52), CKD (*χ*^2^=0.33, p = 0.56), CEVD χ =0.26, p = 0.61), hematologic malignancy (*χ*^2^=2.8, p = 0.1), immune suppressant use (*χ*^2^=0.09, p = 0.77), hypertension (*χ*^2^= 0.22, p = 0.64), alcohol related disorders (*χ*^2^= 0.171, p = 0.68), and MS (*χ*^2^= 2.4, p = 0.12).

**Table 2.** Time-to-Event Analysis: West Nile Neuroinvasive Disease (WNND)

| **Covariate** | **Hazard Ratio** | **95% Confidence Interval** | **P-value** |
| --- | --- | --- | --- |
| <sup>a</sup>Male | 1.290 | (1.148, 1.451) | < 0.001 |
| <sup>a</sup>Age (per 10 years) | 1.105 | (1.062, 1.149) | < 0.001 |
| Ischemic heart diseases | 1.000 | (0.838, 1.193) | 0.996 |
| Other forms of heart disease | 1.141 | (0.989, 1.316) | 0.070 |
| <sup>a</sup>Chronic kidney disease | 1.206 | (1.002, 1.451) | 0.047 |
| <sup>a</sup>Cerebrovascular diseases | 1.224 | (1.031, 1.452) | 0.021 |
| Other chronic obstructive pulmonary disease | 1.086 | (0.782, 1.506) | 0.623 |
| Diabetes mellitus | 1.016 | (0.872, 1.183) | 0.842 |
| <sup>a</sup>Hematologic malignancy | 1.383 | (1.088, 1.758) | 0.008 |
| Black or African American | 1.176 | (0.957, 1.446) | 0.123 |
| Diseases of liver | 0.845 | (0.675, 1.059) | 0.144 |
| Not Hispanic or Latino | 0.947 | (0.835, 1.074) | 0.394 |
| <sup>a</sup>Immune Suppressants | 1.428 | (1.114, 1.830) | 0.005 |
| <sup>a</sup>Essential (primary) hypertension | 1.182 | (1.044, 1.338) | 0.008 |
| <sup>a</sup>Alcohol related disorders | 1.537 | (1.200, 1.970) | 0.001 |
| <sup>a</sup>Multiple sclerosis | 2.338 | (1.622, 3.370) | < 0.001 |

<sup>a</sup>Statistically significant at p<0.05

### Risk factors for mortality

Overall, 13% (392/3064) of patients with WNV infection from 2013 through 2024 died; 33% (129/392) of those deaths occurred within 30 days of WNV diagnosis. Among those who died, 50% (197/392) had a prior diagnosis of WNND. The overall CFR for WNND in this cohort was 16% (197/1206).

Potential risk factors for mortality in the TriNetX cohort that were significant in the bivariate analysis included age, male sex, not Hispanic or Latino, Black or African American, ischemic heart disease, other heart disease, hematologic malignancy, diabetes, HIV, CKD, liver disease, COPD, CEVD, dementia, solid organ transplant, immune suppressants (including subgroups of tacrolimus and MMF), prednisone, and antineoplastics.

In multivariable analysis for mortality, several factors were identified as significant across all timeframes (30 days, 90 days, and overall). For 30-day mortality, these included WNND (aHR 2.49, 95% CI 1.37-4.52), age (per decade) (aHR 1.32, 95% CI 1.07-1.60), other heart disease (aHR 5.50, 95% CI 2.96-10.23), CKD (aHR 2.08, 95% CI 1.01-3.93), and CEVD (aHR 2.00, 95% CI 1.14-3.50) (Table 3). The results for 90-day and overall mortality are available in S Table 4 and S Table 5, respectively. The HRs for WNND, CKD, CEVD, and age were proportional across time windows, while the HR for other heart disease was significantly higher within 30 days. The proportionality assumption was met for WNND (*χ*^2^=0.002, p = 0.96), CKD (*χ*^2^=0.59, p = 0.44), and CEVD (*χ*^2^=1.3, p = 0.26).

**Table 3.** Time-to-Event Analysis: Mortality within 30 days of West Nile Virus infection.

| **Covariate** | **Hazard Ratio** | **95% Confidence Interval** | **P-value** |
| --- | --- | --- | --- |
| <sup>a</sup>WNND | 2.491 | (1.373, 4.522) | 0.003 |
| Male | 0.646 | (0.387, 1.079) | 0.095 |
| <sup>a</sup>Age (per 10 years) | 1.318 | (1.072, 1.598) | 0.008 |
| Ischemic heart diseases | 1.074 | (0.573, 2.011) | 0.824 |
| <sup>a</sup>Other forms of heart disease | 5.499 | (2.955, 10.233) | < 0.001 |
| <sup>a</sup>Chronic kidney disease (CKD) | 2.078 | (1.099, 3.930) | 0.024 |
| <sup>a</sup>Cerebrovascular diseases | 1.996 | (1.137, 3.503) | 0.016 |
| Other chronic obstructive pulmonary disease | 1.504 | (0.661, 3.421) | 0.331 |
| Diabetes mellitus | 0.590 | (0.307, 1.135) | 0.114 |
| Hematologic malignancy | 1.554 | (0.598, 4.040) | 0.366 |
| Black or African American | 0.682 | (0.222, 2.092) | 0.503 |
| Immune Suppressants | 0.581 | (0.165, 2.043) | 0.397 |
| Unspecified dementia | 0.428 | (0.057, 3.210) | 0.409 |
| Diseases of liver | 1.947 | (0.929, 4.081) | 0.078 |
| Human immunodeficiency virus [HIV] disease | 0.572 | (0.062, 5.311) | 0.623 |
| Antineoplastics | 0.715 | (0.094, 5.412) | 0.745 |
| Prednisone | 1.215 | (0.445, 3.318) | 0.704 |
| Not Hispanic or Latino | 0.951 | (0.508, 1.781) | 0.876 |

<sup>a</sup>Statistically significant at p<0.05

## Discussion

Using federated real-world data, we analyzed a large national cohort of patients diagnosed with WNV infection. Our analysis, which to our knowledge is the most extensive contemporary U.S study of WNND and mortality risk factors (over 2,000 patients from 65 HCOs between 2013 and 2024), corroborates previous evidence with a larger sample size and more granularity. Specifically, we identified older age, CKD, and CEVD as risk factors for both WNND and mortality (across all timeframes), and male sex, hematologic malignancy, immune suppressant use, hypertension, alcohol related disorders, and MS as risk factors for WNND.

Our findings are consistent with prior consensus that increased age is associated with WNND and mortality,^8,9^ and male sex has been identified as a risk factor for WNND in roughly half of studies.^8^ Clinical characteristics associated with WNND and mortality in prior studies have varied. A recent meta-analysis identified hypertension, cancer, and diabetes as significantly associated with WNND, while CKD was associated with mortality.^10^ A limited number of studies have identified cardiovascular disease, liver disease, alcohol use, and autoimmune disease as risk factors for WNND and mortality.^8,10^ We identified similar conditions as significant risk factors for WNND; diabetes and liver disease were exceptions, possibly because we did not stratify for disease severity.

Immunocompromising conditions and immunosuppression are also widely considered to be risk factors for WNV disease severity, although most prior analyses have been unable to distinguish between specific conditions and medications owing to sample size limitations.^8,9^ A recent retrospective cohort analysis of patients from the Mayo Clinic found that immunosuppression is associated with severity of clinical manifestations of WNND.^11^ We similarly found that specific immunocompromising conditions (hematologic malignancy and MS) and those receiving immune suppressants (including solid organ transplant recipients) were at increased risk for WNND.

These risk factors align with our current understanding of the pathogenesis of WNND. Multiple components of the immune system (including innate, humoral, and cellular immunity) are involved in mitigating WNV infection.^15^ Additionally, WNV enters the central nervous system (CNS) through multiple pathways, one of which includes permeability of the blood-brain barrier (BBB). ^7,16^ Conditions that disrupt the immune system or cause increased CNS vulnerability through permeability of the BBB or inflammation may lead to increased risk of WNND and sequelae.^15,16^ The risk factors identified in this study for WNND and mortality broadly fit into these two categories – conditions that specifically impair the immune system (hematologic malignancy and immune suppressant use) and conditions that increase CNS vulnerability (hypertension, heart disease, and CEVD). Age, sex, CKD, and MS may contribute to both immune impairment and CNS vulnerability. Further mechanistic and causal inference studies are needed to elucidate the causal pathways by which these risk factors contribute to WNND and mortality.

Overall, we found a greater than two-fold risk of death among those diagnosed with WNND, adjusting for other comorbidities. This risk, coupled with the increasing prevalence of multiple risk factors associated with WNND (such as age, immunocompromise, hypertension, heart disease, CEVD, and CKD) indicates that a growing population in the U.S. is likely at risk for morbidity and mortality following WNV infection.

Our approach of using federated real-world data from TriNetX allowed us to compare multiple characteristics and risk factors across a large cohort. Our study design addressed prior challenges in risk factor analyses for WNV outcomes by including: (1) larger sample sizes to compare multiple risk factors,^9,17^ (2) longer cohort follow-up, given that acute and convalescent phase mortality from WNV may differ, and (3) broader geographic representation.

However, a key limitation to this analysis is that the cases are based on ICD-10 codes. Federated data networks like TriNetX provide the opportunity for analyzing de-identified patient data from multiple regions, providing more statistical power. Nevertheless, local cohorts remain important for further analyzing these observations and exploring specific nuances through access to individual-level data.

Given there is no effective treatment or licensed vaccine for WNV in humans, it is essential to identify populations who are at increased risk for WNV severity. Risk factor analyses could lead to clinical prediction tools for use in triaging patients diagnosed with WNV infection. These analyses could also assist ongoing efforts by public health officials to model and forecast WNND for targeted interventions.^18^ If a WNV vaccine becomes available, a targeted vaccination strategy among those at highest risk for WNND could be cost-effective and impactful.^19^ As the population in the U.S and many countries continues to age with increasing comorbidities, it will be essential to continue to identify and respond to the growing risk of morbidity and mortality from WNV.

## Acknowledgments

This work was supported by the National Institute of Health [T32 AI007291-34] to SDJ and the Fisher Center for Environmental Infectious Diseases. Author contributions: Concept, design, analysis—S.D.J, D.D; Writing—original draft: S.D.J.; Writing—review and editing: S.D.J., D.D.

## Data availability statement

Data for this study were accessed and analyzed using the TriNetX platform.

---

**Figure 1.**
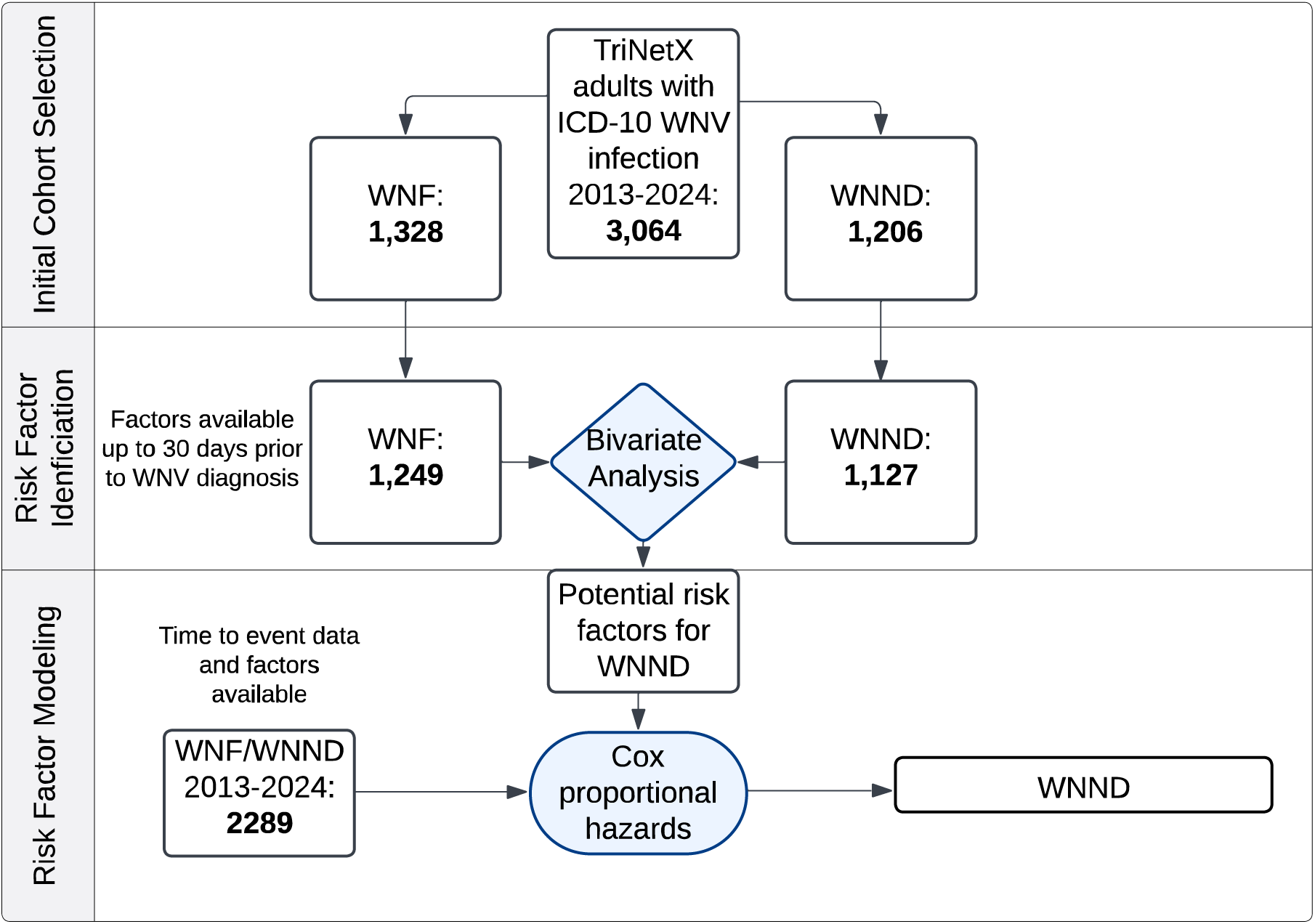
Cohort Selection and Analytic Framework: West Nile Neuroinvasive Disease (WNND) The flow diagram depicts the cohort selection and analysis process in TriNetX. The initial cohort included all patients with ICD-10 codes for WNV infection (with subgroups of WNF and WNND). Those patients with data available within a 30 day-period before or on the same day as WNV diagnosis were included in the bivariate analysis. Significant factors from the bivariate analysis and patients with time-to-event data available were then included in the multivariable Cox proportional hazards model with the outcome of WNND.

**Figure 2.**
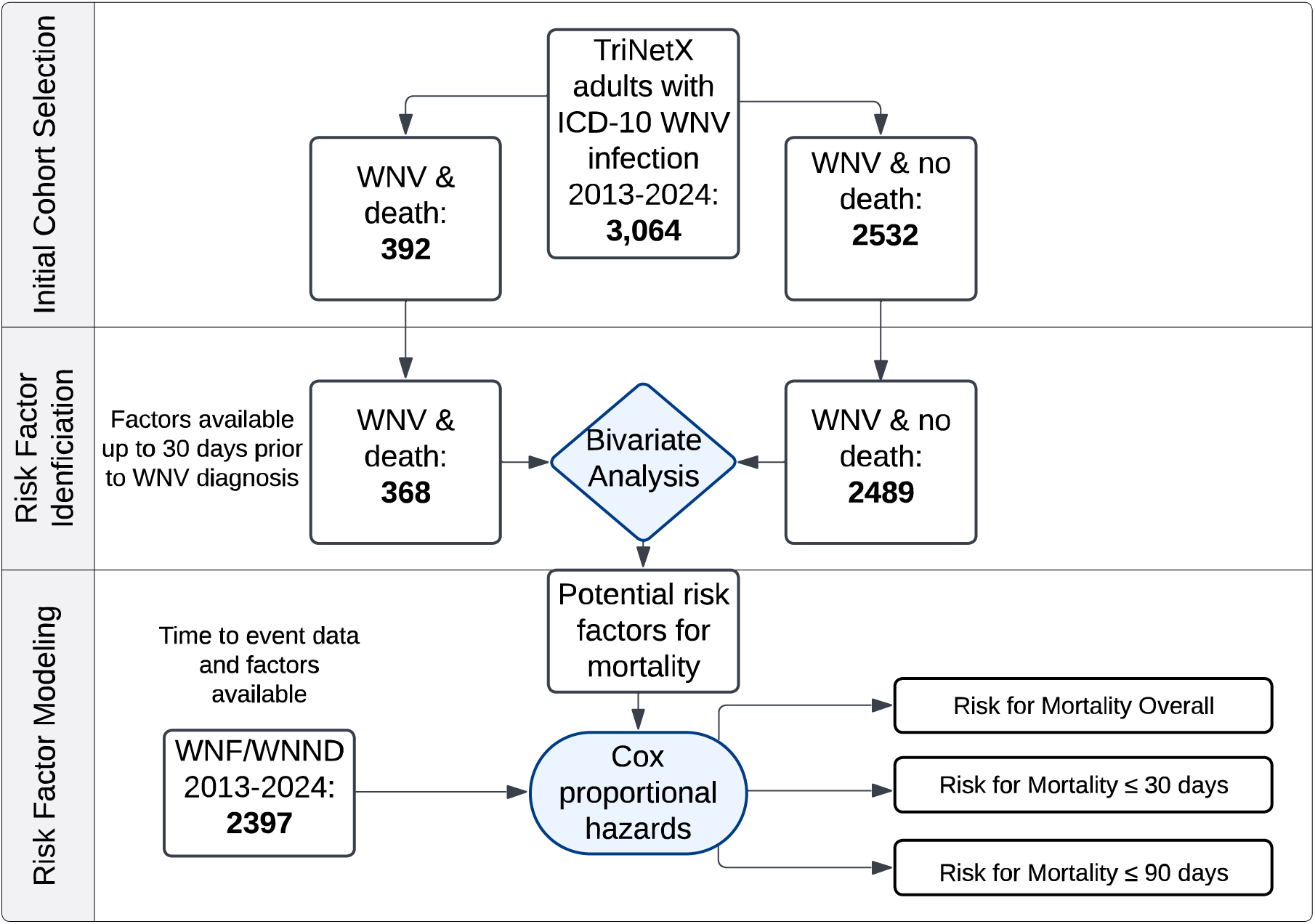
Cohort Selection and Analytic Framework: West Nile Virus (WNV) Mortality. The flow diagram depicts the cohort selection and analysis process in TriNetX. The initial cohort included all patients with ICD-10 codes for WNV infection (with subgroups of including subsequent death or no death). Those patients with data available within a 30 day-period before or on the same day as WNV diagnosis were included in the bivariate analysis. Significant factors from the bivariate analysis and patients with time-to-event data available were then included in multivariable Cox proportional hazards models with the outcomes of mortality overall, mortality within 30 days, and mortality within 90 days.

