## Supplementary Material for "Risk factors for West Nile neuroinvasive disease and mortality in the United States, 2013-2024"

**Supplementary Material: TriNetX Cohort and Analysis**

**S Table 1: TriNetX Cohorts***

| **Cohorts** | **Definition** | **Codes** | **Size** |
| --- | --- | --- | --- |
| WNF Adult ICD-10 2013-2024  (referred to as WNF 2013-2024) | Any adult patient with ICD-10 code indicating West Nile infection without encephalitis or neurologic manifestation from Jan 1^st^ 2013-Dec 31^st^ 2024 | **A92.30**: West Nile virus infection, unspecified,  **A92.39** West Nile virus infection with other complications  **Cannot have:**  **A92.31**: West Nile virus infection with encephalitis  **A92.32**: West Nile virus infection with other neurologic manifestation | **1,328** (60/101 HCOs) |
| WNND Adult ICD-10 2013-2024  (WNND 2013-2024) | Any adult patient with ICD-10 code indicating WNV encephalitis or neurologic manifestation from Jan 1^st^ 2013-Dec 31^st^ 2024 | **A92.31**: West Nile virus infection with encephalitis  **A92.32**: West Nile virus infection with other neurologic manifestation  **Cannot have:**  **A92.30**: West Nile virus infection, unspecified,  **A92.39** West Nile virus infection with other complications | **1,206** (57/101 HCOs) |
| WNV Adult ICD-10 2013-2024 deaths  (WNV & death 2013-2024) | Any adult patient with diagnosed West Nile infection who died from Jan 1^st^ 2013-Dec 31^st^ 2024 | **A92.3**: West Nile virus infection,  **A92.30**: West Nile virus infection, unspecified  **A92.31**: West Nile virus infection with encephalitis  **A92.32**: West Nile virus infection with other neurologic manifestation  **A92.39**: West Nile virus infection with other complications  **AND Deceased** | **392** (44/101 HCOs) |
| WNV Adult ICD-10 2013-2024 non-deceased  (WNV & no death 2013-2024) | Any adult patient with diagnosed West Nile infection from Jan 1^st^ 2013-Dec 31^st^ 2024, excluding deceased | **A92.3**: West Nile virus infection,  **A92.30**: West Nile virus infection, unspecified  **A92.31**: West Nile virus infection with encephalitis  **A92.32**: West Nile virus infection with other neurologic manifestation  **A92.39**: West Nile virus infection with other complications  **Cannot have: Deceased** | **2,532** (65/101 HCOs) |
| WNV Adult all ICD-10 2013-2024  (WNV 2013-2024) | Any adult patient with diagnosed West Nile infection from Jan 1^st^ 2013-Dec 31^st^ 2024 | **A92.3**: West Nile virus infection,  **A92.30**: West Nile virus infection, unspecified  **A92.31**: West Nile virus infection with encephalitis  **A92.32**: West Nile virus infection with other neurologic manifestation  **A92.39**: West Nile virus infection with other complications | **3,064** (65/101 HCOs) |

*Cohort queries made on March 28^th^, 2025

**Risk factor selection**

We first compared baseline characteristics and statistics between cohorts stratified by WNND and mortality. We selected characteristics based on risk factors that previously have been assessed or associated with WNND, severity, and/or mortality from literature review. Among those risk factors, we included those with sample size >10 individuals (which is the minimum sample size number that TriNetX reports to maintain anonymization). To distinguish among categories of immunosuppression, we separated HIV, solid organ transplant, and immune suppressant medications into separate categories. For cancer we specified hematologic malignancies, as this was the largest ICD-10 group in TriNetX that did not include benign neoplasms.

Our time window for risk factors included up to 30 days prior to and including the day of the index event (diagnosis of WNV infection). We selected among risk factors that were significant in bivariate analysis (p <0.05) to use for the multivariable analyses.

**S Table 2. Initial Cohort Characteristics Assessed**

| Covariate | TriNetX Criteria |
| --- | --- |
| Age | Age at index (WNV diagnosis) |
| Sex | Male, Female, Unknown |
| Ethnic group | Hispanic or Latino, Not Hispanic or Latino, Unknown |
| Race | White, Black or African American, Asian, Native Hawaiian or Other Pacific Islander, American Indian or Alaska Native, Other, Unknown |
| Malignant neoplasms of lymphoid, hematopoietic and related tissue (Hematologic malignancy) | C81-C96 |
| Ischemic heart disease | I20-I25 |
| Other forms of heart disease | I30-I5A |
| Diabetes | E08-E13 |
| HIV | B20 |
| CKD | N18 |
| Liver Disease | K70-74 |
| Alcohol related disorders | F10 |
| Multiple Sclerosis | G35 |
| Dementia | F03 |
| Asthma | J45 |
| COPD | J44 |
| Essential (primary) hypertension | I10 |
| Cerebrovascular disease | I60-163 |
| Rheumatoid Arthritis | M06 |
| Solid Organ Transplant | Z94 |
| Antineoplastics | AN00 |
| Immune suppressants | IM600 (includes calcineurin inhibitors, biologics) |
| Prednisone | 8640 |
| Tacrolimus | 42316 |
| Mycophenolate mofetil | 68149 |

| **S Table 3. Baseline Characteristics of WNV & death vs WNV & no death 2013-2024 Cohorts** | | | | | | |
| --- | --- | --- | --- | --- | --- | --- |
| **Covariate** | | **Patients, No. (%)** | | | | **^a^P-value** |
|  |  | **WNV & death**  **(n = 368)** | | **WNV & no death**  **(n = 2489)** | |  |
| Age at diagnosis, mean (SD), y | | 69 (13) | | 56 (7) | | <0.0001 |
| Sex | |  | |  | |  |
| Female | | 117 (32) | | 1099 (44) | | <0.0001 |
| Male | | 243 (66) | | 1349 (54) | | <0.0001 |
| Unknown | | ^b^**≤**10 (3) | | 41 (2) | | 0.15 |
| Ethnicity | |  | |  | |  |
| Not Hispanic or Latino | | 305 (83) | | 1892 (76) | | 0.004 |
| Hispanic or Latino | | 23 (6) | | 198 (8) | | 0.25 |
| Unknown ethnicity | | 40 (11) | | 399 (16) | | 0.01 |
| Race | |  | |  | |  |
| White | | 270 (73) | | 1924 (77) | | 0.1 |
| Black or African American | | 41 (11) | | 166 (7) | | 0.002 |
| Asian | | 13 (4) | | 55 (2) | | 0.12 |
| Other Race | | ^b^**≤**10 (3) | | 123 (5) | | 0.06 |
| Native Hawaiian or Other Pacific Islander | | ^b^**≤**10 (1) | | ^b^**≤**10 (1) | | 0.19 |
| American Indian or Alaska Native | | ^b^**≤**10 (1) | | ^b^**≤**10 (1) | | 0.19 |
| Unknown Race | | 29 (8) | | 204 (8) | | 0.83 |
| Hematologic malignancy | | 28 (8) | | 76 (3) | | <0.0001 |
| Ischemic heart diseases | | 78 (21) | | 201 (8) | | <0.0001 |
| Other forms of heart disease | | 148 (40) | | 464 (19) | | <0.0001 |
| Diabetes mellitus | | 74 (20) | | 352 (14) | | 0.003 |
| HIV | | ^b^**≤**10 (3) | | 21 (1) | | 0.001 |
| Chronic kidney disease | | 70 (19) | | 170 (7) | | <0.0001 |
| Diseases of Liver | | 36 (10) | | 140 (6) | | 0.002 |
| Essential (primary) hypertension | | 112 (30) | | 745 (30) | | 0.84 |
| Alcohol related disorders | | 17 (5) | | 86 (3) | | 0.26 |
| Cerebrovascular diseases | | 70 (19) | | 213 (9) | | <0.0001 |
| Chronic obstructive pulmonary disease | | 29 (8) | | 74 (3) | | <0.0001 |
| Asthma | | 16 (4) | | 115 (5) | | 0.81 |
| Multiple sclerosis | | ^b^**≤**10 (3) | | 36 (2) | | 0.07 |
| Dementia | | 12 (3) | | 29 (1) | | 0.002 |
| Other rheumatoid arthritis | | 11 (3) | | 47 (2) | | 0.16 |
| Transplanted organ | | 17 (5) | | 55 (2) | | 0.006 |
| Immune suppressants (overall) | | 27 (7) | | 67 (3) | | <0.0001 |
| Tacrolimus | | 13 (4) | | 42 (2) | | 0.02 |
| Mycophenolate | | 16 (4) | | 30 (1) | | <0.0001 |
| Antineoplastics | | 15 (4) | | 56 (2) | | 0.04 |
| Prednisone | | 42 (11) | | 156 (6) | | 0.0003 |
| **^a^**For continuous data calculated using independent t-tests and categorical data using χ^2^ or Fisher’s exact tests  ^b^TriNetX patient counts **≤** 10 are rounded up to 10 to maintain anonymity | | | | | | |

**Multivariable analysis**

We compared the association between selected risk factors that were statistically significant in the bivariate analysis (p <0.05 in S Table 3) in multivariable Cox proportional hazards models. Covariates were measured within 30 days prior to and including the day of the index event (diagnosis of WNV infection). Mortality was assessed at 30 days, 90 days, and any time after the index event. For the outcome of WNND, this was assessed any time after the index event, and the cohort included individuals with a healthcare visit in 2013-2024 prior to WNV diagnosis. Given the overlap between immune suppressant use and solid organ transplant, we selected immune suppressant use as a covariate for our final models given the larger sample size.

**Multivariable Cox Proportional Hazards Models: Mortality Outcomes**

| **S Table 4. Mortality within 90 days** | | | |
| --- | --- | --- | --- |
| **Covariate** | **Hazard Ratio** | **95% Confidence Interval** | **P-value** |
| ^a^WNND | 2.580 | (1.578, 4.218) | < 0.001 |
| Male | 0.858 | (0.558, 1.319) | 0.485 |
| ^a^Age (per 10 years) | 1.280 | (1.094, 1.508) | 0.003 |
| Ischemic heart diseases | 1.088 | (0.646, 1.833) | 0.750 |
| ^a^Other forms of heart disease | 3.940 | (2.401, 6.465) | < 0.001 |
| ^a^Chronic kidney disease | 2.235 | (1.332, 3.752) | 0.002 |
| ^a^Cerebrovascular diseases | 1.892 | (1.167, 3.067) | 0.010 |
| Other chronic obstructive pulmonary disease | 1.048 | (0.489, 2.242) | 0.905 |
| Diabetes mellitus | 0.727 | (0.434, 1.220) | 0.228 |
| Hematologic malignancy | 1.889 | (0.904, 3.947) | 0.091 |
| Black or African American | 0.874 | (0.386, 1.983) | 0.748 |
| Immune Suppressants | 1.095 | (0.499, 2.405) | 0.821 |
| Unspecified dementia | 0.498 | (0.115, 2.154) | 0.351 |
| ^a^Diseases of liver | 2.142 | (1.179, 3.891) | 0.012 |
| HIV | 0.752 | (0.151, 3.757) | 0.728 |
| Antineoplastics | 1.661 | (0.568, 4.858) | 0.354 |
| Prednisone | 1.585 | (0.776, 3.236) | 0.206 |
| Not Hispanic or Latino | 0.866 | (0.518, 1.446) | 0.582 |

^a^Statistically significant at p<0.05

| **S Table 5. Overall Mortality** | | | |
| --- | --- | --- | --- |
| **Covariate** | **Hazard Ratio** | **95% Confidence Interval** | **P-value** |
| ^a^WNND | 2.001 | (1.556, 2.572) | < 0.001 |
| Male | 1.281 | (0.997, 1.647) | 0.053 |
| ^a^Age (per 10 years) | 1.509 | (1.370, 1.660) | < 0.001 |
| Ischemic heart diseases | 1.359 | (0.999, 1.849) | 0.051 |
| ^a^Other forms of heart disease | 1.716 | (1.302, 2.261) | < 0.001 |
| ^a^Chronic kidney disease | 1.992 | (1.450, 2.737) | < 0.001 |
| ^a^Cerebrovascular diseases | 1.411 | (1.041, 1.914) | 0.027 |
| ^a^Other chronic obstructive pulmonary disease | 1.764 | (1.159, 2.686) | 0.008 |
| Diabetes mellitus | 1.245 | (0.941, 1.646) | 0.125 |
| ^a^Hematologic malignancy | 1.619 | (1.009, 2.597) | 0.046 |
| Black or African American | 1.508 | (0.993, 2.288) | 0.054 |
| Immune Suppressants | 1.330 | (0.830, 2.131) | 0.236 |
| Unspecified dementia | 1.029 | (0.509, 2.080) | 0.937 |
| ^a^Diseases of liver | 2.056 | (1.410, 2.998) | < 0.001 |
| HIV | 0.695 | (0.247, 1.953) | 0.490 |
| ^a^Antineoplastics | 1.758 | (1.013, 3.052) | 0.045 |
| ^a^Prednisone | 1.856 | (1.232, 2.798) | 0.003 |
| Not Hispanic or Latino | 1.268 | (0.916, 1.755) | 0.153 |
| ^a^Statistically significant at p<0.05 | | | |
